# TRANSLATION AND VALIDATION OF A SPANISH PRELIMINARY VERSION OF THE PACKA QUESTIONNAIRE AMONG PATIENTS WITH FIBROMYALGIA

**DOI:** 10.64898/2026.01.03.26343375

**Authors:** Klara Albajes, Mayte Serrat, Albert Feliu-Soler, G. Lorimer Moseley, Miriam Almirall, Roland R. Reezigt

## Abstract

**Background:** Patients’ understanding of pain and its biopsychosocial nature is thought to be important in order to optimize treatment decisions and treatment outcomes. A validated Spanish questionnaire for patients based on a biopsychosocial model assessing knowledge and attitudes of pain was unavailable. The aim of this study is to validate the Spanish version of the PACKA, to have a Spanish questionnaire to be used among pain patients in both clinical and research settings.

**Methods:** Phase 1: forward and backward translation, and expert committee; Phase 2: a cross-sectional and longitudinal study among Fibromyalgia patients. In the cross-sectional study (n =330), internal consistency, structural validity and hypothesis testing were examined.

**Results:** The 26-item PACKA was translated and validated in 4 stages. Acceptable internal consistency and test-retest reliability was demonstrated. Internal consistency: Cronbach’s α =0.776. Hypotheses testing: associations with the Neurophysiology of Pain Questionnaire r =0.31.

**Conclusion:** This Spanish version of PACKA has shown acceptable validity and reliability in patients with Fibromyalgia. Further use and reanalysis of the questionnaire might improve this preliminary validation.

## 1. INTRODUCTION

In the last decades, the understanding of patients’ pain has gained in relevance and there is increasing emphasis on providing a contemporary view of pain based on the biopsychosocial model in which all factors, biological, psychological and social, play an important role (Louw et al., 2016; Ryan et al., 2010; Nijs et al., 2014; Foster et al., 2003; Butler and Moseley, 2013). This is due to the large body of evidence suggesting that patient’s knowledge related to *all* physiological and processing mechanisms involved in pain phenomena can contribute to improved self-reported intervention options, self-management and self-control (Moseley and Butler, 2015) and better health outcomes (Camerini et al., 2013; Musekamp et al., 2019). Therefore, the long-standing demand to address patients’ beliefs and attitudes about their pain and its treatment (Schwartz et al., 1985) finally seems to be taking a more prominent role.

Being able to capture how someone understands pain and its underlying biopsychosociality offers many clinical and research benefits. From a clinical point of view, it may allow to optimise and increase knowledge to the extent necessary and accurate to promote the most optimal individual and personal treatment decisions. From a research point of view, it can help us understand the relationship between cognitive and behavioural processes and how they contribute to individuals’ health.

There are available questionnaires that aim to assess knowledge, attitudes and beliefs of practitioners (Beetsma et al, 2020; Bishop et al., 2010; Darlow et al., 2014; Domenech et al., 2013; Houben et al., 2004; Moran et al., 2017; Pincus et al., 2012; Waddell et al., 1993). The latest questionnaire Knowledge and Attitudes of Pain (KNAP) (Beetsma et al., 2020) was developed to assess both biopsychosocial attitudes towards pain and knowledge about modern pain science among health practitioners. However, there are fewer tools specifically for patients (Reezigt & Beetsma et al., 2025).

With this purpose, a new questionnaire, Pain, Attitudes, Cognitions and Knowledge Assessment (PACKA) was based on the KNAP and on extensive work with recovered consumers to identify what concepts they consider most important for enabling their recovery (Leake et al., 2021; Moseley et al., 2014). The PACKA was developed specifically for people with pain and to capture both cognitive aspects, such as knowledge, and behavioural aspects, such as attitudes.

This study aimed to 1) translate the first version of the PACKA questionnaire into Spanish and 2) to test psychometric properties (i.e. structural and criterion validity) in a Spanish sample of patients with fibromyalgia. Additional correlations (e.g., with the HADS) could have been examined; however, given the preliminary nature of this version, the validation was intentionally limited to structural and criterion validity. Extending the analysis to emotional constructs was considered premature at this early stage of questionnaire development.

## 2. METHODS

The study used a two-phase approach to translate and validate the preliminary version of the PACKA.

### 2.1 Phase I – Translation into Spanish and face validity

Following the recommendations for translating questionnaires (Beaton et al., 2000), four stages were used: forward translation, backward translation, expert committee, and preliminary pilot testing. First, the initial translation from English to Spanish was made by a native Spanish speaker and pain specialist with proficiency in English. As a specialist, the translator took into account the concepts that the questionnaire is intended to measure in order to be as faithful as possible to the original questionnaire. Secondly, the translation was translated back into English by a native English translator with proficiency in Spanish. As recommended in the guidelines (Beaton et al., 2000) this second translator did not know the concepts of the questionnaire to avoid bias. Third, two independent pain specialists formed the expert committee and reviewed both versions, resolved discrepancies and reached a consensus on all items to generate the final version of the translated questionnaire. Fourth and last, face validity was tested on a small sample of 10 people diagnosed also with fibromyalgia.

### 2.2 Phase II - Testing measurement properties

#### 2.2.1. Design and participants

For structural validity, data from a cross-sectional survey among patients with Fibromyalgia, obtained as part of another study (Serrat et al., 2022), were used. All participants were consecutively registered in the Vall d’Hebron University Hospital - Central Sensitivity Syndromes Specialised Unit and were evaluated by a rheumatology clinician and a physical therapist to guarantee the selection criteria.

The inclusion criteria were 18–75 years of age, fulfill the FM classification criteria based on 2010/2011 American College of Rheumatology (Wolfe et al., 2010), and be able to understand Spanish. Individuals who had participated in concurrent or past randomised controlled trials (RCTs) (throughout the previous year) were excluded, as were individuals suffering any comorbid condition such as severe mental health disorders or neurodegenerative diseases. The study was conducted in the context of regular clinical practice at the Vall d’Hebron University Hospital. Before the survey, all participants signed an informed consent form.

Sociodemographic information (age, gender, and educational level) had been previously collected and reported in an earlier study and was used in the present analysis to describe the characteristics of the sample (Serrat et al., 2022). For the present validation, two independent cohorts from that same study were used: one cohort was used to evaluate structural validity, and a second independent cohort was used to test criterion validity. Both cohorts followed the same recruitment procedures and were obtained from the Central Sensitivity Syndromes Specialized Unit at Vall d’Hebron University Hospital.

#### 2.2.2 Measures

##### 2.2.2.1. Primary measure

Preliminary PACKA questionnaire. This tool was developed by Beetsma et al as adaptation of the KNAP, and encompasses 26 items used to assess knowledge, beliefs and attitudes about pain. The PACKA scoring system is not based on correct versus incorrect responses, but on the extent to which each response reflects a biopsychosocial (BPS) or biomedical (BM) orientation. Each item is anchored in one of these two conceptual perspectives. Answers aligned with the BPS interpretation receive a +1; answers reflecting a BM interpretation receive a –1. An “I don’t know” response is scored as 0, as it does not indicate a clear orientation toward either perspective. Consequently, total scores can range from –26 to +26, with higher scores reflecting a stronger BPS orientation and lower scores indicating a more biomedical perspective.

##### 2.2.2.2. Secondary measures

The Revised Neurophysiology of Pain Questionnaire (NPQ) in Spanish (Torres-Lacomba et al., 2021). The NPQ contains 13 ‘true’, ‘false’, ‘unsure’ items relating to the neurophysiology of pain (Catley et al., 2013). The total score ranges from 0 to 13, with higher scores indicating more correct responses. The Spanish NPQ contains a good level of reliability (ICC 0.90) and internal consistency (Cronbach’s α =0.82) for evaluating the knowledge of the neurophysiology of pain among patients (Torres-Lacomba et al., 2021).

##### 2.2.2.3. Data analysis

All study outcomes were analyzed with descriptive statistics for both samples used and expressed as median (IQR) for most quantitative variables, and percentages (%) and frequencies for categorical variables.

Structural validity was tested using an exploratory factor analysis. Sampling adequacy was analyzed with the Kaiser-Meyer-Olkin and Bartlett’s test. Construct validity was done with an exploratory factor analysis, using a promax rotation. The number of factors was chosen based upon the scree plot, simulated parallel analysis and underlying rationale. In addition, factor retention and item adequacy were evaluated using standard EFA criteria, including minimum primary loadings of ≥ 0.40, absence of substantial cross-loadings (≥ 0.30–0.40), acceptable uniqueness values (≤ 0.40), and the proportion of variance explained by each factor, ensuring that the retained factors contributed meaningfully to the overall solution. Further, internal consistency was tested using Cronbach’s α. Last, criterion validity was tested based on an expected correlation with the NPQ, where only knowledge was tested, which was a priori set as r = 0.3 due to the partial overlap.

## 3. RESULTS

### 3.1. Phase I - Translation into Spanish

In the first stage, the 26-item PACKA questionnaire was translated into Spanish. Following this, the translated and Spanish version of PACKA was translated back to English and this was compared to the original version with a satisfactory result. In the second stage, both experts from the expert committee, after reaching a consensus, generated the Spanish version of PACKA which was approved in the final stage (Fig. 1).

**Figure 1.**
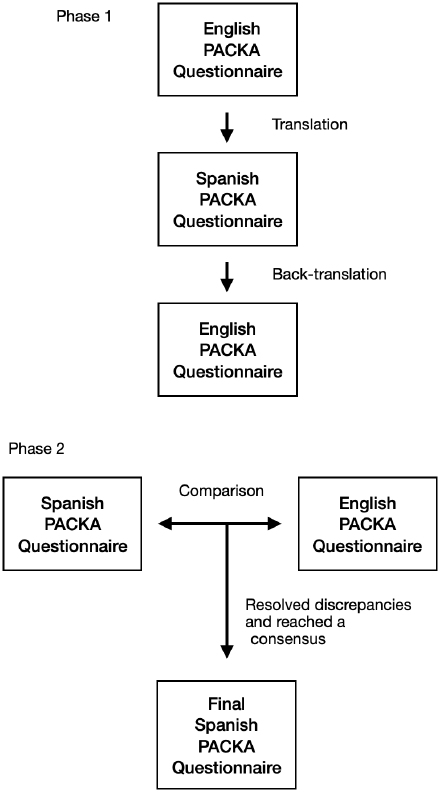
Translation and cross-cultural adaptation process of the PACKA questionnaire.

### 3.2. Phase II - Measurement properties

#### 3.2.1. Sociodemographic and clinical characteristics of the study samples

Two samples were used for the structural and criterion validity. Both were comparable in composition, mostly females in their fifties as shown in Table 1.

**Table 1.**
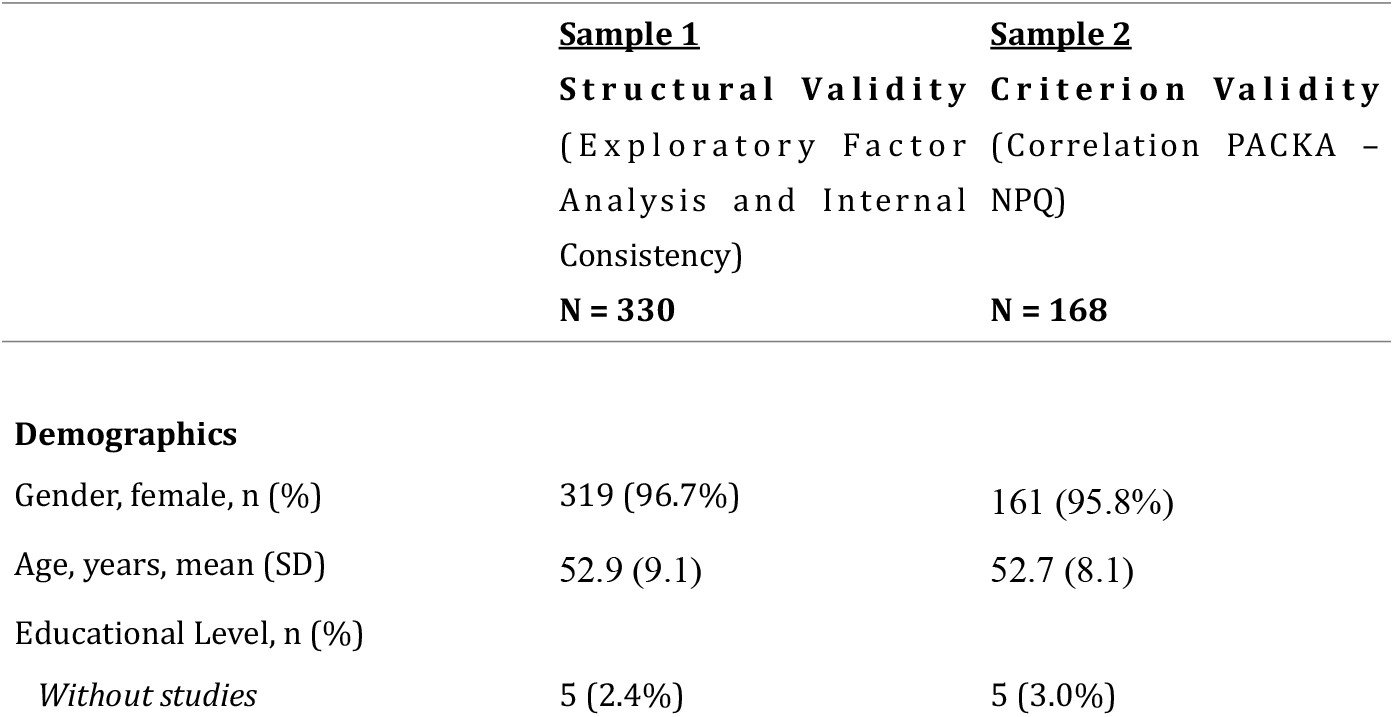
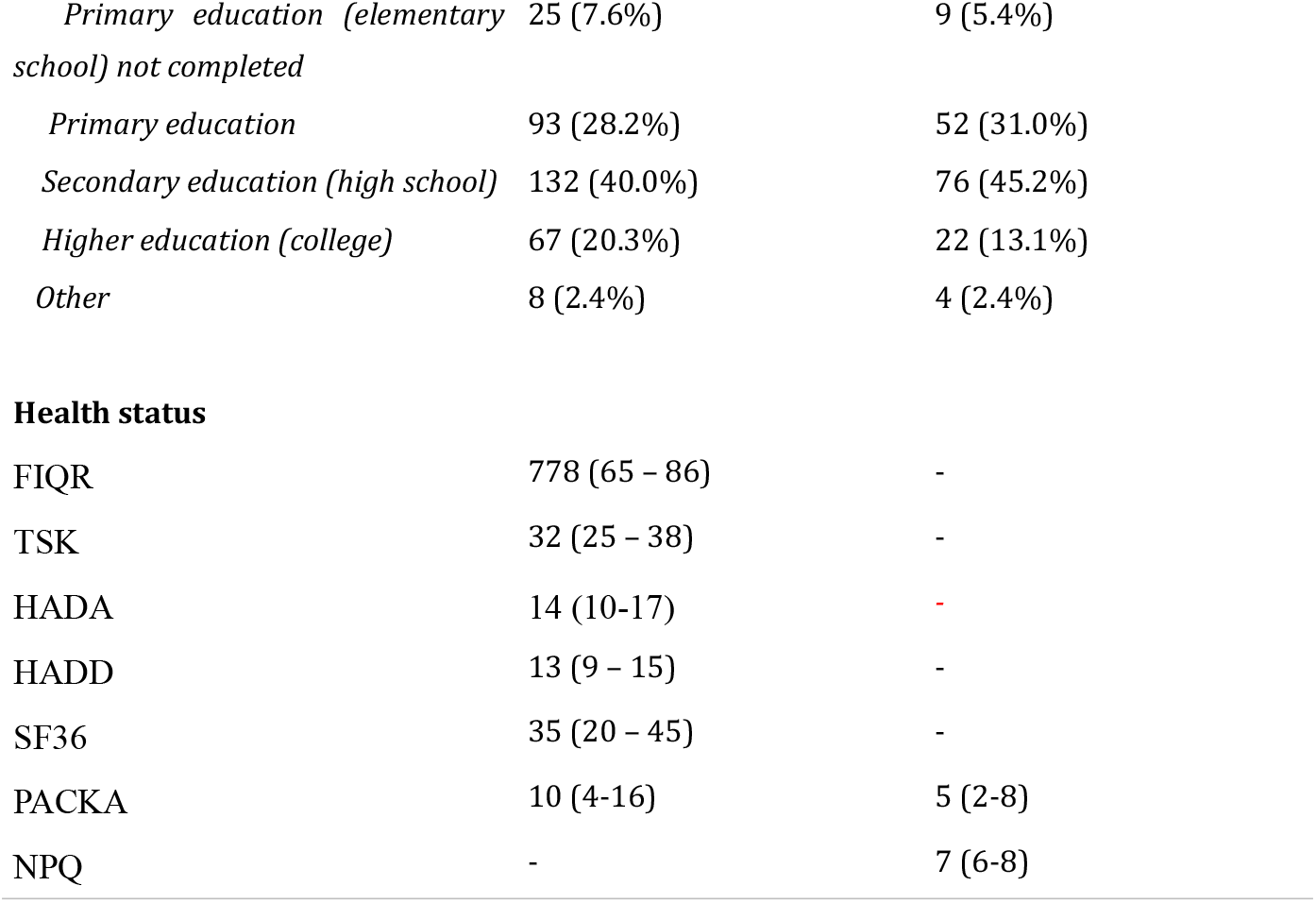
Characteristics of the study samples. All data is shown as median (IQR), unless otherwise specified. Note: FIQR: Revised Fibromyalgia Impact Questionnaire; TSK: Tampa Scale for Kinesiophobia; HADA: Hospital Anxiety and Depression Scale - Anxiety; HADD: Hospital Anxiety and Depression Scale - Depression; SF36: Physical Functioning component of the 36-Item Short Form Survey; PACKA: Pain, Attitudes, Cognitions and Knowledge Assessment; NPQ: Neurophysiology of Pain Questionnaire.

PACKA score distributions for both samples are presented in Figure 2, showing that the two cohorts performed similarly on the questionnaire.

**Figure 2.**
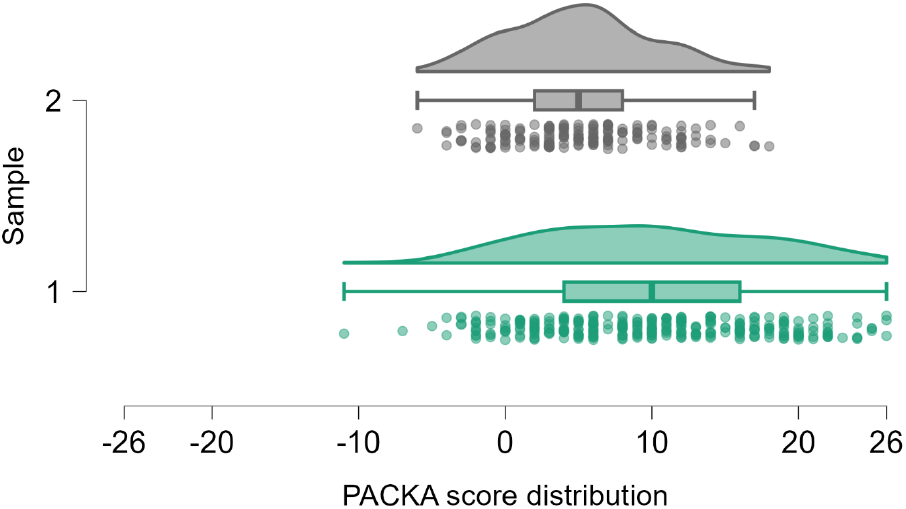
Distribution of the PACKA scores in both samples

#### 3.2.2. Structural Validity

The Kaiser–Meyer–Olkin measure confirmed sampling adequacy (KMO = 0.817), and Bartlett’s test of sphericity was significant (χ^2^(324) = 1836.086, p <.001).

The EFA revealed a three-factor structure, which was interpreted as 1) Understanding the multifactorial nature of pain, 2) Perception of one’s own ability to manage pain and 3) Understanding the underlying contributors to persistent pain. Importantly, the three-factor solution was consistent with the factor structure identified in the original Dutch version of the PACKA. Items loading on each factor, as well as cross-loadings and uniqueness values, are presented in Table 2.

**Table 2.**
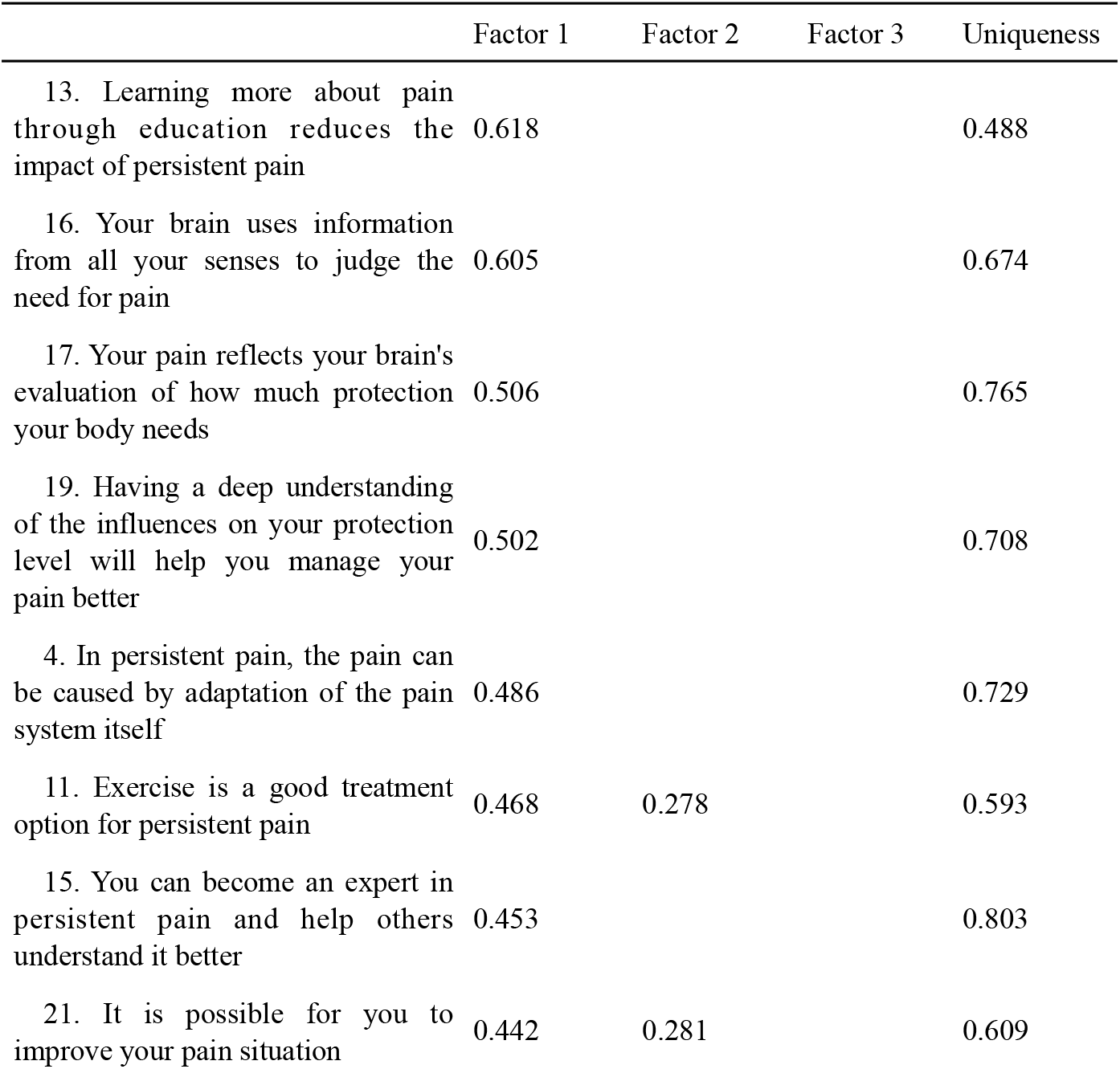
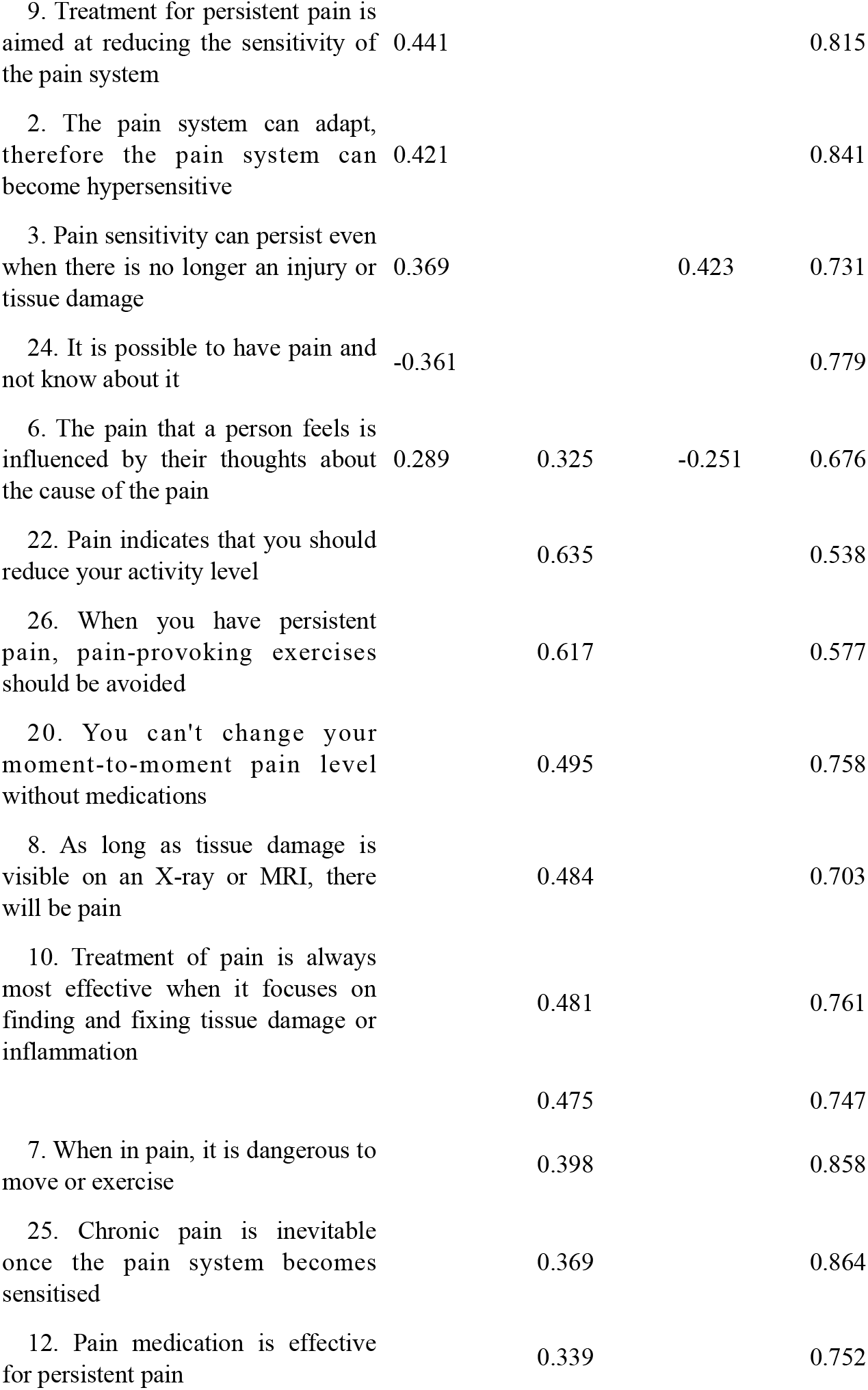
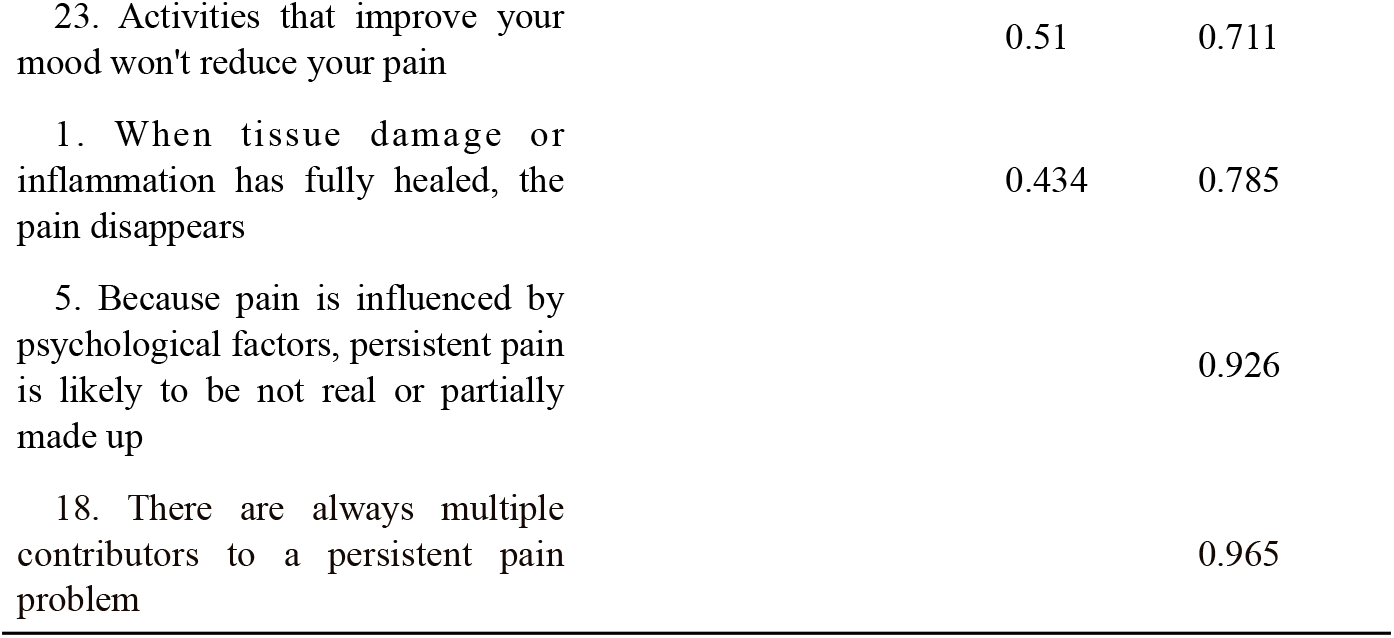
Factor loadings and uniqueness values for the 26-item PACKA questionnaire. *Note*. Sorted on factor loading. Applied rotation method is promax

*Factor Loadings and Uniqueness in the Exploratory Factor Analysis*

#### 3.2.3. Internal consistency

The overall internal reliability was Cronbach’s α=0.776 (95% CI: 0.741 – 0.804). None of the items were of strong enough influence to be further examined.

#### 3.2.4. Criterion Validity testing

Correlation between PACKA and NPQ was: r =0.313 (95% CI 0.167; 0.458), p <.001, confirming the hypotheses, thereby supporting construct validity (see Figure 3 for the correlation scatterplot and dispersion graphs).

**Figure 3.**
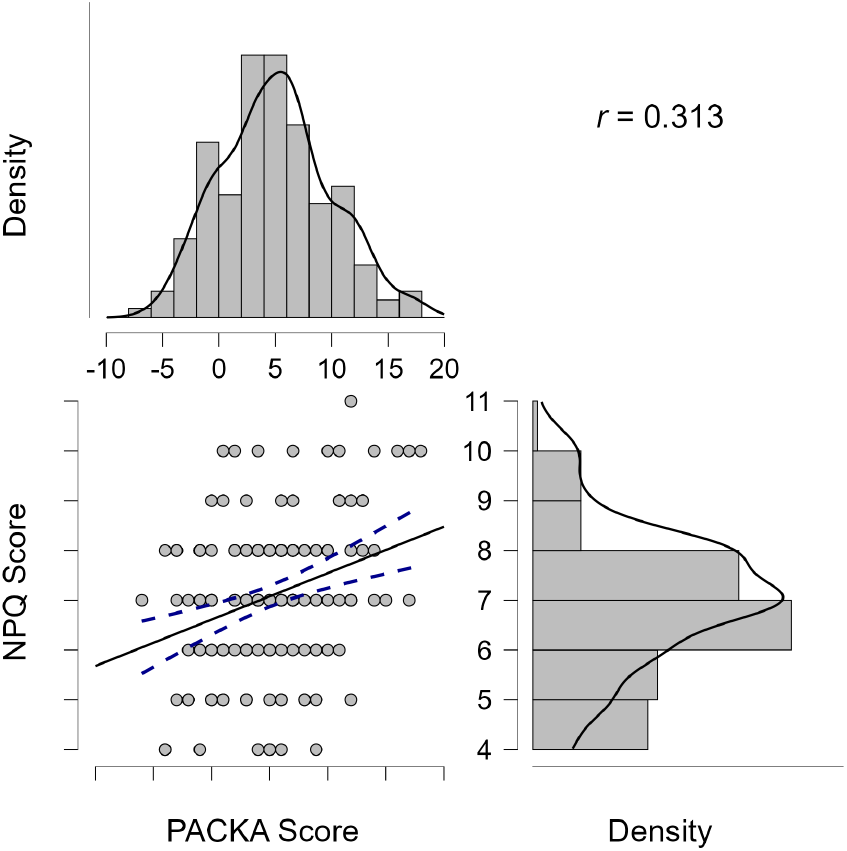
Correlation scatterplot including dispersion graphs

## 4. DISCUSSION AND CONCLUSIONS

The aim of this study was to translate the PACKA questionnaire into Spanish and to examine its preliminary psychometric properties in a sample of patients with fibromyalgia. The results indicate that the Spanish version of the PACKA shows acceptable preliminary measurement properties: the exploratory factor analysis supported a three-factor structure that is consistent with the factor solution identified in the original Dutch version, internal consistency was adequate, and the expected association with the NPQ was observed. These findings suggest that the questionnaire captures relevant aspects of pain-related knowledge and beliefs within a biopsychosocial framework.

The questionnaire demonstrates adequate psychometric properties for its use. The internal validity and reliability suggest that Spanish PACKA has sufficient sensitivity to differentiate different levels of biopsychosocial pain knowledge consistently. This tool allows assessment of patients’ knowledge, beliefs and attitudes about pain, particularly as it relates to a more biomedical or a biopsychosocial conception. In clinical settings, this questionnaire can detect understandings, beliefs and attitudes that are inconsistent with contemporary scientific understanding and/or likely to be unhelpful for recovery. This can also guide educational intervention on a personalised level. It is feasible that PACKA score may have utility in establishing a cut-off point of necessary knowledge to start an interdisciplinary treatment program based on biopsychosocial approach to treatment. From program developers’ perspectives, such insight would optimize time and resources. In the research field, this tool may contribute to deeper understanding of the interrelationships that occur between different cognitive-behavioural processes related to pain, e.g. anxiety, catastrophising and knowledge in psychological-related factors in pain.

The psychometric properties between NPQ and PACKA are moderate. The mere fact of this result between the two tools indicates a relationship between the content measured. However, the correlation was low. Three considerations are relevant here. First, the NPQ was first developed by one individual with lived experience of chronic pain and recovery, and advanced understanding of pain-related neurophysiology. The NPQ did not consider patient perspectives, nor employ Item Response Theory or other recognised methods to develop assessment tools. This may result in inappropriate terminology, too sophisticated concepts, or suboptimal item fit (although the NPQ 13 item tool, which resulted from Rasch analysis of the original 21 item tool, contains items with satisfactory fit). Relevant here is the conclusion from that Rasch study, that this tool could be improved by using adapted language (e.g. metaphors) rather than concepts that are the hardest to grasp (Catley et al., 2013). Second, the NPQ was based on understanding of the neurophysiology of pain at the end of the last century – that scientific understanding has progressed since then raises the possibility that it may reflect outdated understandings. Third, the two questionnaires are targeting mildly different things: while the NPQ might be more biomedical factors orientated (Catley et al., 2013), the PACKA questionnaire has been developed specifically in order to discriminate the biopsychosocial knowledge of pain. That is to say, the PACKA questionnaire encompasses knowledge, beliefs, and attitudes, which the NPQ does not.

A final consideration of the current work relates to the preliminary nature of the face validity assessment conducted in Phase I. The qualitative evaluation was performed on a very small and homogeneous group of participants, which limits the representativeness of this initial appraisal and may constrain the content validity of the Spanish version. To strengthen the robustness of the instrument, future studies should evaluate face validity in larger and more diverse samples, including individuals with different socio-educational backgrounds, ages, and clinical profiles.

In summary, the Spanish version of PACKA is a useful tool for assessing patients’ pain-related beliefs and attitudes. These results suggest adequate psychometric properties for its use with fibromyalgia patients. Nevertheless, authors believe that measurement properties can be improved by expanding both the psychometric analysis and the target population. Also, the use of the questionnaire may provide new information for future analyses.

## Data Availability

The data that support the findings of this study are available from the corresponding author upon reasonable request. Due to the sensitive nature of clinical data, the data are not publicly available.

